# Tract-based Quantitative MRI for Resolving the Clinico-Radiological Paradox in Multiple Sclerosis

**DOI:** 10.1101/2025.09.17.25335756

**Authors:** Osama Abdullah, Puti Wen, Abdelmalek W. Abdelrazeq, Mariya Matrosova, Lev Brylev, Amar Ahmad, Vasiliy Bryukhov, David Melcher, Bas Rokers

## Abstract

The clinico-radiological paradox in multiple sclerosis (MS) arises because conventional MRI measures, particularly total lesion volume, fail to fully capture the true burden of disability.

These broad volumetric measures overlook the dual influence of where lesions occur and what their microstructural composition is. This study aimed to improve the clinical relevance of lesion analysis in MS by combining tract-based spatial filtering with quantitative microstructural MRI metrics. We hypothesized that filtering lesions through clusters of functionally meaningful white matter tracts, rather than broad anatomical compartments, would enable more accurate identification of clinically relevant damage.

We studied 132 participants, including 89 patients with MS (49 relapsing–remitting, 17 primary progressive, 23 secondary progressive; 62 women and 27 men) and 43 healthy controls (28 women and 15 men). All underwent standardised 3 Tesla MRI including FLAIR, T1 mapping, magnetisation transfer ratio (MTR), and diffusion tensor imaging (DTI) with fractional anisotropy (FA) and mean diffusivity (MD). We evaluated associations between imaging measures and disability and assessed predictive performance with ridge-penalised regression across EDSS, motor scores (T25FW, 9HPT), and MSPro-defined progression risk.

Tract-based models remained superior to classical region-based models, achieving higher discrimination and better model fit for binarised EDSS and MSPro (AUC = 0.86–0.96 vs 0.57– 0.86) and substantially greater variance explained for continuous motor outcomes (RZ = 0.245– 0.43 vs 0.01–0.195).

By integrating lesion location and microstructural composition, tract-based quantitative MRI enhances disability prediction and provides interpretable imaging markers to support disability characterisation and individualized monitoring in MS.

**Abbreviated Summary:** Abdullah et al. show that anchoring lesions within functionally critical white-matter tracts and measuring their microstructural tissue composition reveals stronger associations with motor and progression-related clinical measures in multiple sclerosis, helping explain why conventional lesion burden alone often poorly reflects disability.

## Introduction

Multiple sclerosis (MS) is a chronic, immune-mediated disease of the central nervous system characterized by inflammatory demyelination and neurodegeneration, most prominently in white matter, with post-mortem studies showing that lesions vary widely in demyelination, axonal loss, gliosis, and microglial activity depending on stage, location, and patient-specific factors. ^1–3^ This heterogeneity is mirrored clinically in the diverse disability profiles seen in patients, ranging from gait and motor impairment to visual dysfunction, cognitive decline, and sensory symptoms, raising the challenge of how lesion characteristics can be meaningfully linked to functional outcome.^4–8^ Conventional MRI, particularly T2 and Fluid-Attenuated Inversion Recovery (FLAIR), guides diagnosis and monitoring but depicts lesions with varying signal intensities across broad anatomical regions, so lesion counts and volumes often miss true disease burden. This shortcoming gives rise to the clinico-radiological paradox, a bidirectional disconnect where high lesion burden may result in minimal impairment, or conversely, small lesions in strategic locations may cause prominent disability, underscoring the need for more advanced imaging approaches that can bridge this disconnect.^9–11^

Quantitative MRI (qMRI) offers promising complementary measures that augment conventional MRI by probing tissue microstructure beyond visible lesions. Metrics such as magnetization transfer ratio (MTR) and T1 relaxation times are sensitive to microstructural changes associated with myelin and axonal content, although they lack pathological specificity.^12,13^ MTR and T1 metrics have been used to characterize tissue integrity within lesions, successfully stratifying them by disease severity and stage.^14,15^ A particularly compelling finding came from a longitudinal study showing that reductions in MTR could be detected in normal-appearing white matter (NAWM) up to four months before new lesions became visible on conventional MRI.^16^ Diffusion tensor imaging (DTI) probes tissue microstructure by characterizing water diffusion. Scalar metrics such as reduced fractional anisotropy (FA) and increased mean (MD) or radial diffusivity (RD) can reflect axonal disorganization and demyelination, even in NAWM, notwithstanding the influence of other factors such as inflammation or edema on these indices.^17^ By reconstructing white matter tracts from diffusion imaging, studies have moved beyond gross anatomical compartments, providing a more precise mapping of disease impact and revealing correlations with clinical disability than region-based approaches.^18–27^ Together, these findings highlight the potential of qMRI and tract-based approaches to capture clinically meaningful pathology that conventional lesion measures overlook.

Parallel to imaging advances, clinical outcomes are now assessed with multiple domain-specific scales: the Expanded Disability Status Scale (EDSS, global disability scale weighted toward ambulation and motor function at higher scores),^28^ Symbol Digit Modalities Test (SDMT, for processing speed),^29^ Timed 25-Foot Walk (T25FW, for gait),^30^ Nine-Hole Peg Test (9HPT, for manual dexterity),^31^ and MSPro scale (for progression risk).^32,33^ While these developments enrich both imaging and disability assessment, most previous studies relied on simple univariate correlations, often using a single clinical score. Moreover, previous studies frequently used gross anatomical compartments to explain disability (for example, correlating EDSS with MTR in periventricular lesions; *r* = 0.316, *p* = 0.007),^34^ an approach that, although informative, lacks functional specificity and reinforces oversimplified interpretations of “global disability.” This reliance on univariate and regionally crude analyses introduces confounds, observer bias, and noise, thereby perpetuating the paradox.

In this study, we propose a tract-aware, multiparametric framework that integrates qMRI metrics with the functional anatomy of white matter tracts. By quantifying both lesion location (where) and microstructural composition (what), and correlating these measures with multiple disability scores across progressively structured statistical models, we aim to establish stronger and more specific structure-function relationships. We hypothesize that this comprehensive approach will better account for lesion heterogeneity and the multidimensional nature of disability, enabling improved patient stratification and more accurate distinction of MS phenotypes compared to conventional lesion volume measures.

## Materials and Methods

### Subject Selection, Clinical Assessment, and Data Management

A total of 132 participants (89 MS patients and 43 healthy controls) were recruited between June 2022 and June 2024 at the Research Center of Neurology in Moscow, Russia. Consent was obtained in accordance with the Declaration of Helsinki. The study was approved by the Local Ethics Committee of the Research Center of Neurology (Moscow, Russia).

MS patients were clinically classified to relapsing remitting (RRMS), primary progressive (PPMS), or secondary progressive (SPMS) subtypes based on the 2017 Revised McDonald Criteria,^35^ with inclusion limited to individuals aged 18–70. Control subjects, aged 18–70, were included if they had no cognitive or neurological disorders, and no structural brain abnormalities on MRI beyond age-related nonspecific white matter hyperintensities. General exclusion criteria for all participants included MRI contraindications (e.g., pacemaker), pregnancy, recent corticosteroid therapy (within 30 days for MS patients), structural brain pathology, or corrupted MRI data. All subjects provided written informed consent prior to participation. Clinical disability was assessed using the EDSS, T25FW, 9HPT (dominant and non-dominant hand), and MSPro. We focused our analysis on EDSS ( a global disability scale that, particularly at higher scores, is strongly weighted toward ambulation and motor function) together with MSPro, T25FW, and 9HPT, which emphasize motor and physical disability domains with overlapping functional substrates. The SDMT was also administered to all participants, including healthy controls, and is reported descriptively in Table 1 to characterise the cohort, but SDMT was not used as an outcome in any of the imaging–disability models in this study; dedicated SDMT-based analyses, including the corresponding cognitive networks, are reserved for a separate investigation with a larger cohort. Table 1 summarizes the demographic characteristics, Fig.1 summarizes the clinical profiles, and Supplementary Table 1 compares the five disability assessment tools used in this study.

**Figure 1.**
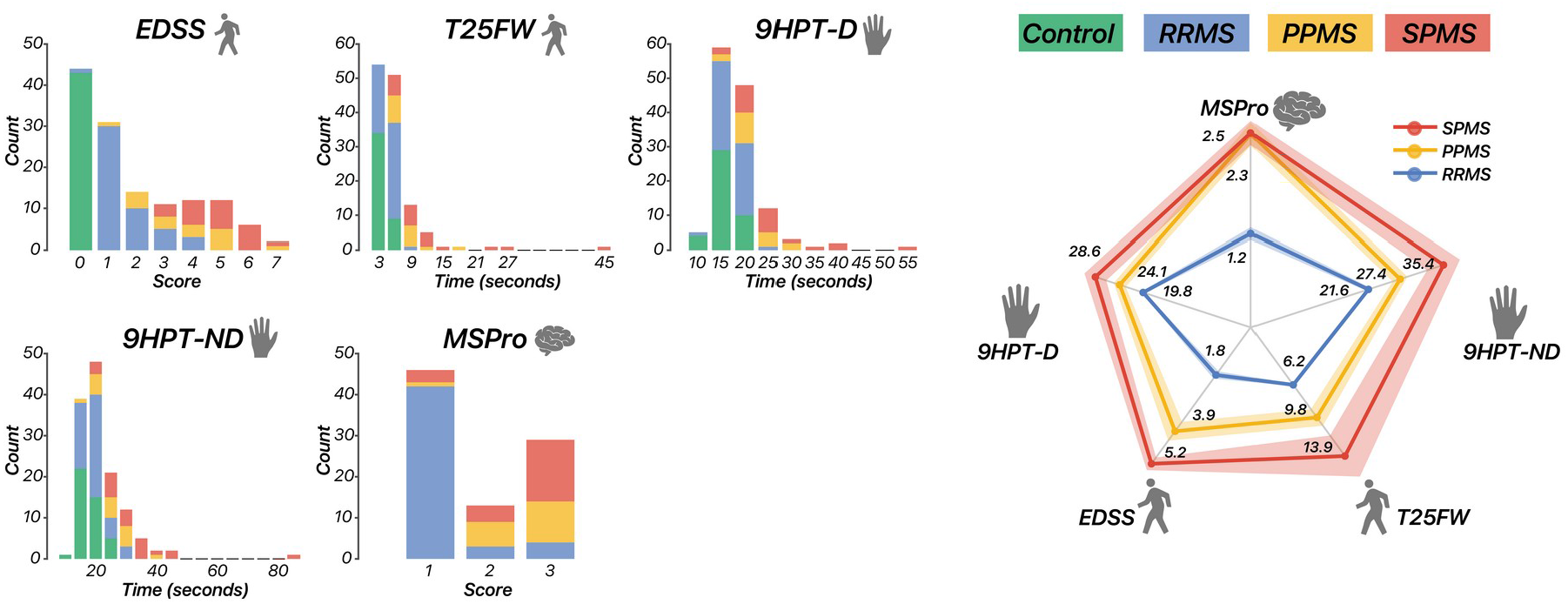
Cohort clinical score distributions across MS subtypes. The radar plot (right) displays group-wise mean values ± standard error of the mean (SEM) for five clinical scores (EDSS, 9HPT-D, 9HPT-ND, T25FW, and MSPro) across RRMS (n = 49), PPMS (n = 17), and SPMS (n = 23) groups. Bar plots (left) represent the same metrics shown individually with their corresponding numerical scales; healthy controls (n = 43) are included where applicable. Data are presented as mean ± SEM. Descriptive statistics for these clinical scores are summarised in Table 1.

**Table 1.** Demographic and clinical characteristics of study participants. Data are presented as mean ± standard deviation (SD) for continuous variables, median [interquartile range (IQR)] for ordinal variables (EDSS, MSPro), and n (%) for categorical variables. Groups: healthy controls ( n = 43), RRMS (n = 49), PPMS (n = 17), SPMS (n = 23). Each participant represents one independent experimental unit.

| Parameter | Healthy Controls | RRMS | PPMS | SPMS |
| --- | --- | --- | --- | --- |
| N (% Female) | 43 (65%) | 49 (76%) | 17 (59%) | 23 (65%) |
| Age | 35.60 ± 10.83 | 35.65 ± 10.25 | 47.88 ± 7.01 | 48.87 ± 12.50 |
| Disease duration (years) | - | 9.17 ± 7.48 | 7.53 ± 5.26 | 17.22 ± 8.37 |
| EDSS | - | 1.5 [1.0-2.0] | 4.0 [2.5-5.0] | 5.5 [4.5-6.0] |
| Therapy: 1st line | - | 29 (59.2%) | 2 (11.8%) | 10 (43.5%) |
| Therapy: 2nd line | - | 15 (30.6%) | 7 (41.2%) | 6 (26.1%) |
| T25FW (s) | 5.63 ± 0.52 | 6.23 ± 1.01 | 9.76 ± 3.57 | 13.95 ± 9.94 |
| 9HPT-D (s) | 18.47 ± 2.31 | 19.81 ± 2.65 | 24.12 ± 4.51 | 28.61 ± 9.37 |
| 9HPT-ND (s) | 20.00 ± 2.87 | 21.56 ± 3.89 | 27.39 ± 5.58 | 35.39 ± 14.16 |
| SDMT Total | 61.88 ± 10.24 | 56.49 ± 11.03 | 46.88 ± 8.78 | 38.43 ± 8.07 |
| MSPro | - | 1.0 [1.0-1.0] | 3.0 [2.0-3.0] | 3.0 [2.0-3.0] |

### MRI Acquisition

A multiparametric 3T MRI protocol was acquired on a Siemens Magnetom Prisma (Syngo MR VE11) at the Research Center of Neurology in Moscow, using a 64-channel Head/Neck coil. FLAIR imaging was acquired using a 3D T2-weighted SPACE sequence with the following parameters: TR = 7000 ms, TE = 390 ms, TI = 2200 ms, flip angle = 120°, voxel size = 0.47 × 0.47 × 0.60 mm3, the sequence was acquired at a 256 × 256 matrix and reconstructed to 512 × 512 using scanner in-plane interpolation. T1-mapping was performed using a 3D MP2RAGE sequence with TR = 5000 ms, TE = 2.98 ms, TI1 = 700 ms, TI2 = 2500 ms, flip angles = 4° and 5° for the first and second inversions respectively, and isotropic voxel size = 1.00 × 1.00 × 1.00 mm3, matrix size = 256 × 240, FOV = 256 × 240 mmZ. Magnetization transfer imaging was acquired using a 3D gradient-echo sequence with TR = 34 ms, TE = 6 ms, flip angle = 10°, voxel size = 1.00 × 1.00 × 3.00 mm3, matrix size = 256 × 192, FOV = 256 × 192 mmZ. Both MT-on and MT-off images were collected for MTR calculation. Diffusion-weighted imaging was performed using a single-shot echo-planar imaging sequence with TR = 5600 ms, TE = 82 ms, flip angle = 90°, matrix size = 122 × 122, multiband acceleration factor = 2. A multi-shell diffusion scheme was employed with b-values of 0, 1000, and 2500 s/mmZ, using 64 diffusion directions per shell. To correct for susceptibility-induced geometric distortions, a reversed phase encoding b=0 acquisition was acquired as well. The total acquisition time was approximately one hour.

### Image Preprocessing

Anatomical segmentation utilized the UNI image from the MP2RAGE acquisition after noise removal with the PreSurfer toolkit (https://github.com/srikash/presurfer). Cortical and subcortical structures were reconstructed using FreeSurfer (v7.4.1, recon-all) and FLAIR images were registered to the structural space.^36^ Lesions were segmented with a custom-trained U-Net implemented in MONAI and manually corrected in FSLeyes by a physician (AWA), with verification by a trained neurologist (LB).^37,38^ Quantitative parameter mapping was then performed using qMRLab,^39^ which provides validated implementations of quantitative MRI models, including the MP2RAGE signal model for estimating T1 relaxation times from the dual-inversion-time acquisition. T1 relaxation maps were estimated from the MP2RAGE sequence with the mp2rage model (TI1/TI2 = 700/2500 ms, flip angles = 4°/5°, TR = 5000 ms). Magnetization transfer ratio (MTR) maps were computed from MT-on and MT-off images using the formula: MTR = [(S₀ − Smt)/S₀] × 100%, where S₀ and Smt represent signal without and with MT saturation, respectively. Diffusion preprocessing was carried out with TractoFlow (Nextflow pipeline) in the Scilus 1.5.0 Singularity container including susceptibility distortion correction (TOPUP) and eddy current correction (EDDY) in FSL.^40^ DTI reconstruction yielded FA and MD maps. FA and MD maps were derived exclusively from the b=1000 s/mmZ shell, as the TractoFlow pipeline restricts FSL’s dtifit to single-shell data for DTI fitting. Local deterministic tractography was performed in DSI Studio with FA-based seeding (threshold = 0.1). Preprocessed DWI, b-values, and b-vectors were converted into diffusion source (.src) files, followed by generalized Q-sampling imaging (GQI; sampling length ratio = 1.25) aligned to the T1 space. Using the resulting fiber orientation distribution (.fib) files, 24 canonical white matter tracts were automatically reconstructed with DSI Studio’s ICBM-152 template-based deterministic tracking.^41^ All imaging metrics (FLAIR, T1 map, MTR, FA, MD) and tract masks were co-registered to the individual T1 structural space for consistency in subsequent analyses (Fig. 2). Co-registration was performed using FreeSurfer’s mri_coreg (rigid-body transformation), followed by mri_vol2vol for resampling to native T1 space.

**Figure 2.**
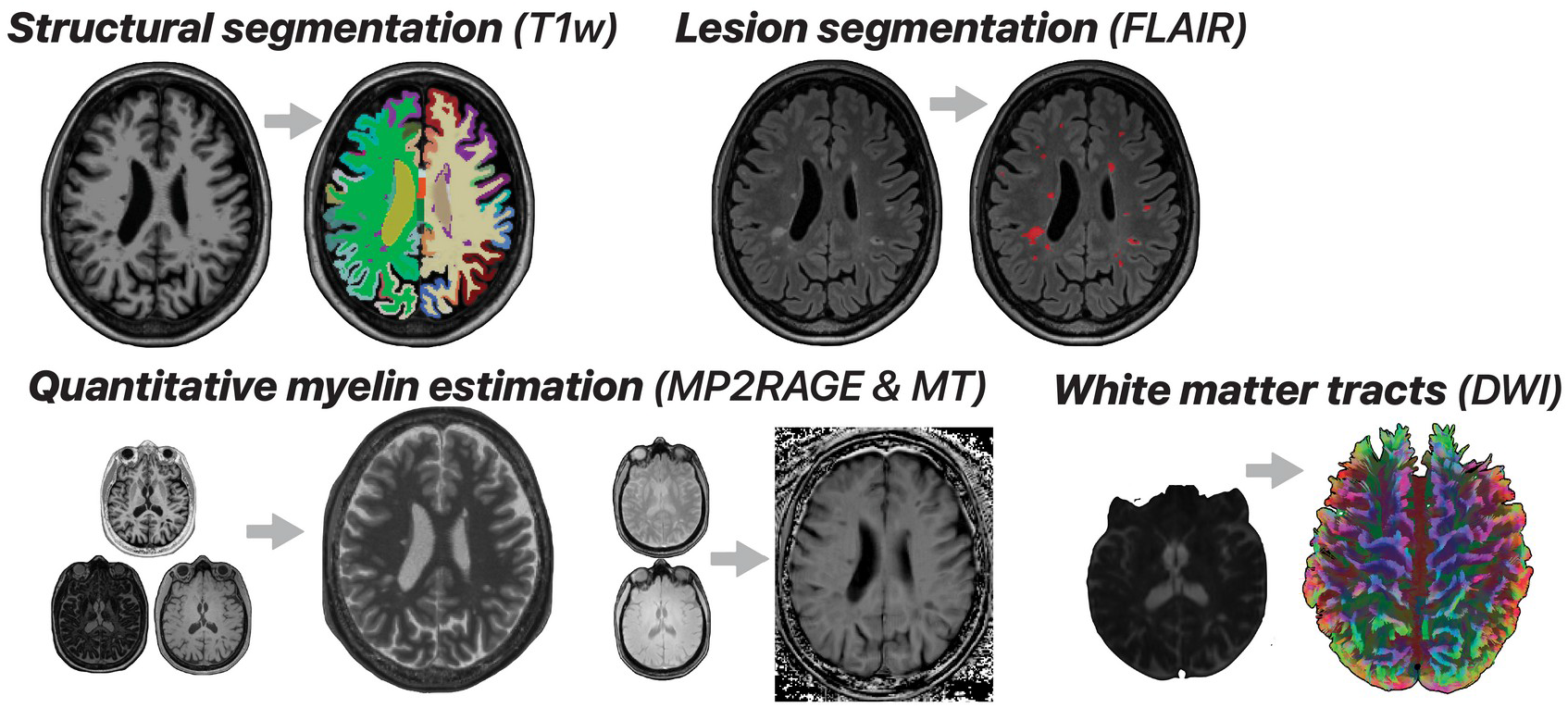
MRI acquisition and preprocessing pipeline. UNI images from the MP2RAGE sequence (TR = 5000 ms, TI1/TI2 = 700/2500 ms, flip angles = 4°/5°, isotropic voxel = 1.0 mm3) were used for anatomical segmentation in FreeSurfer (v7.4.1). FLAIR scans (256×256 matrix, reconstructed to 512×512 via scanner in-plane interpolation) provided lesion masks generated by a custom-trained U-Net model with manual correction. Quantitative maps included T1 relaxation time (estimated via qMRLab MP2RAGE signal model) and magnetization transfer ratio (MTR = [(S₀ − Sₘₜ)/S₀] × 100%). Diffusion preprocessing (TractoFlow, Scilus 1.5.0) included susceptibility distortion correction (FSL TOPUP using a reversed phase-encoding b = 0 acquisition) and eddy current correction (FSL EDDY); FA and MD maps were derived from the b = 1000 s/mmZ shell only.

### Lesion Load and Quantitative Metrics

#### Lesion location metrics

Lesion quantification was performed using two complementary frameworks: a classical anatomical definition, familiar to neurologists (periventricular, juxtacortical, infratentorial, and deep white matter), and a tract-based definition grounded in functional white matter pathways. In the classical framework, lesion masks were first intersected with the four canonical anatomical compartments, and lesion load was recorded as (1) lesion number (LN, count of discrete lesions), (2) lesion volume (LV, cumulative lesion volume in ml), and (3) normalized lesion volume (Lnorm, LV expressed as a percentage of total intracranial volume). In the tract-based framework, 24 subject-specific white matter fiber pathways which were collapsed into four functional clusters (association, projection-brainstem, occipitoparietal, and cerebellar pathways). For each cluster, lesion load was likewise defined as LN, LV, and Lnorm (LV expressed as a percentage of tract volume). Specific tract assignments are listed in Supplementary Table 2 with representative renderings in Fig. 3. Where lesions overlapped multiple tracts (confluent lesions), shared voxels were retained in all intersecting tract-specific lesion maps, reflecting the multi-tract involvement of such lesions. This dual-scale (classical- based vs tract-based) framework enabled direct testing of whether pathway-specific lesion mapping offers greater clinical insight than conventional anatomical compartmentalization.

**Figure 3.**
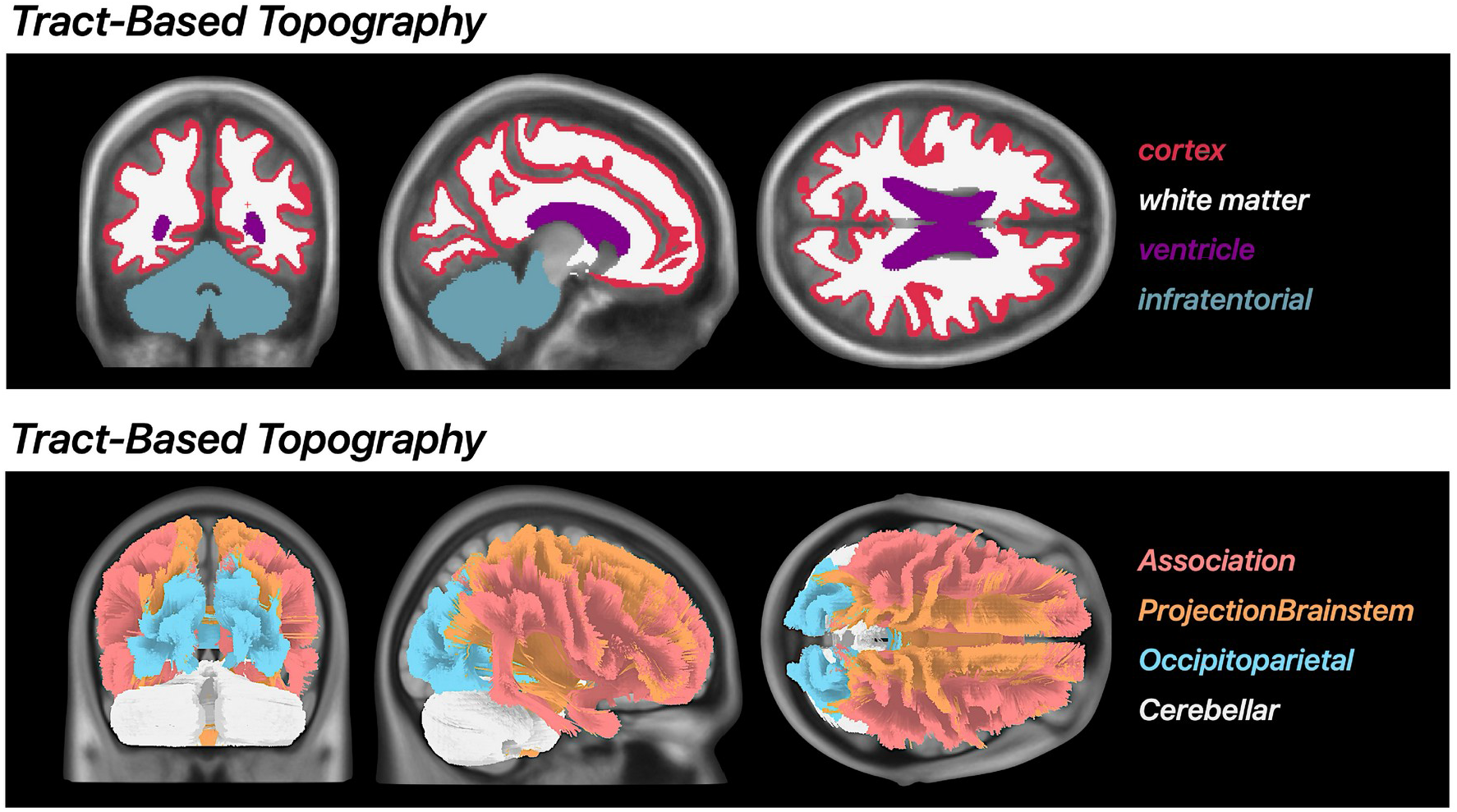
Defining lesion locations. Lesions were classified using two complementary frameworks: (top panel) classical anatomical regions (periventricular, juxtacortical, infratentorial, deep white matter) and (bottom panel) tract-based regions corresponding to 24 canonical white matter pathways collapsed into four functional clusters (association, projection-brainstem, occipitoparietal, cerebellar). Four representative topographies are shown for comparability.

#### Lesion composition metrics

For each participant, mean T1, MTR, FA, and MD values were calculated within lesions falling inside (1) the four classical compartments and (2) the four tract-based clusters. Because tract clusters form clearly defined fiber bundles, the same metrics were also sampled from NAWM by excluding lesion voxels from tract masks, yielding an internal reference free of visible pathology on 3D FLAIR. This generated, for each subject, a matrix of eight lesion-level measurements (four metrics × two location schemes) and a set of tract-specific NAWM values. NAWM sampling was not attempted for the classical compartments, whose diffuse anatomical borders preclude a reliable lesion-free reference. The combined lesion-based and tract-NAWM metrics constituted the lesion-composition feature set supplied to the statistical modeling stage.

### Statistical Analysis

#### Descriptive Group Summaries

The objective was to explore subtype separation patterns by visualizing group-wise mean values and standard errors of the mean (SEM) of quantitative MRI metrics (T1 relaxation, MTR, FA, and MD) across MS subtypes, stratified by both tract-based lesion location and lesion tissue composition (including NAWM), using polar (radar) plots. Figures 1, 4, and 5 present descriptive group-level summaries; no inferential between-group testing was performed on these figures, as the study’s inferential framework is multivariable predictive modelling (reported in Table 2, Figure 6, and Supplementary Tables 3–4) rather than univariate subtype comparison.

**Figure 4.**
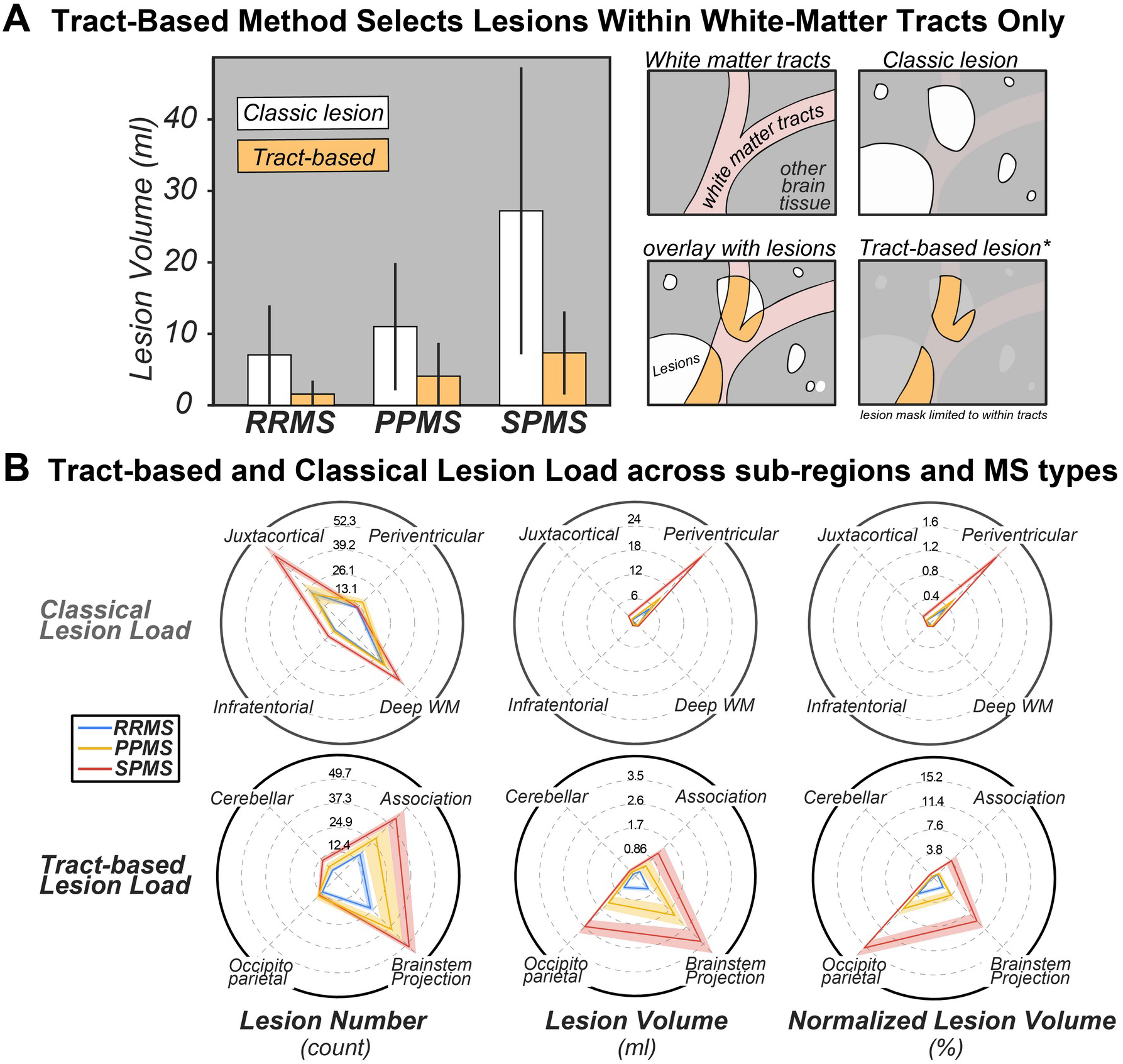
Group-wise comparison of lesion load frameworks across MS subtypes. (**A**) *Left:* Bar plots of absolute whole-brain lesion volumes (mL) for classical (white bars) and tract- based (orange bars) lesion definitions across RRMS (n = 49), PPMS (n = 17), and SPMS (n = 23); error bars represent SEM across subjects within each subtype. The classical definition treats each lesion as a single contiguous mask without anatomical constraint, whereas the tract- based definition retains only lesion voxels intersecting predefined white-matter tracts. *Right:* Schematic of the tract-based filtering mechanism; only voxels falling within white-matter tracts (or tract-lesion intersections) are retained, excluding non-tract lesion extent. (**B**) Radar plots comparing lesion number (count), lesion volume (mL), and normalized lesion volume (%) across MS subtypes (RRMS: blue; PPMS: yellow; SPMS: red), shown for classical anatomical regions (top) and tract-based regions (bottom). Note that the regions plotted differ between rows. Values represent group means ± SEM.

**Figure 5.**
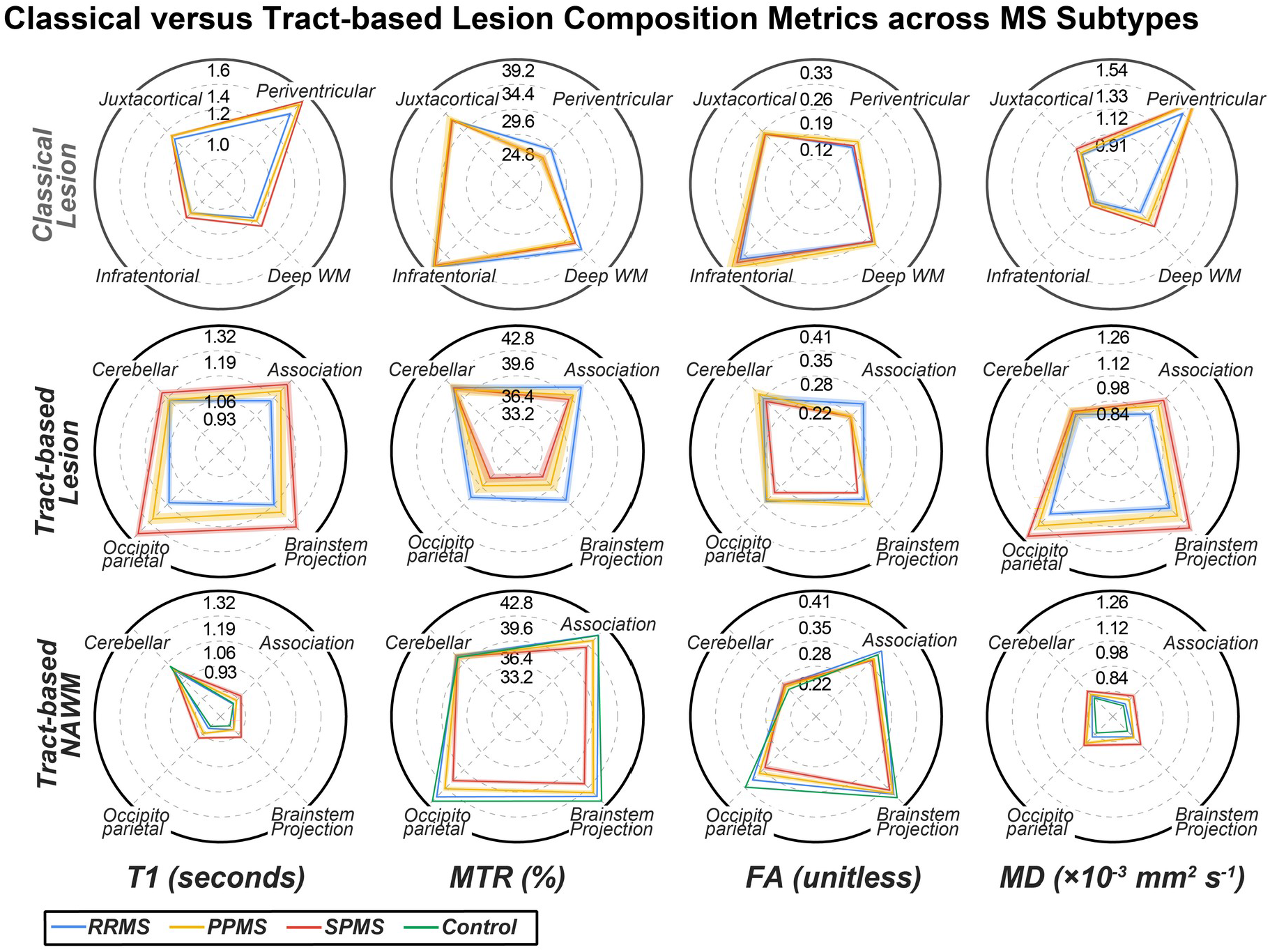
Radar plots of lesion composition metrics across MS subtypes and healthy controls. Quantitative MRI metrics (T1 relaxation time [s], MTR [%], FA [unitless], MD [×10⁻3 mmZ/s]) are shown for classical lesions (top row), tract-based lesions (middle row), and tract-based NAWM (bottom row) across RRMS (n = 49; blue), PPMS (n = 17; yellow), SPMS (n = 23; red), and healthy controls (n = 43; green). All values are group means ± SEM.

**Figure 6.**
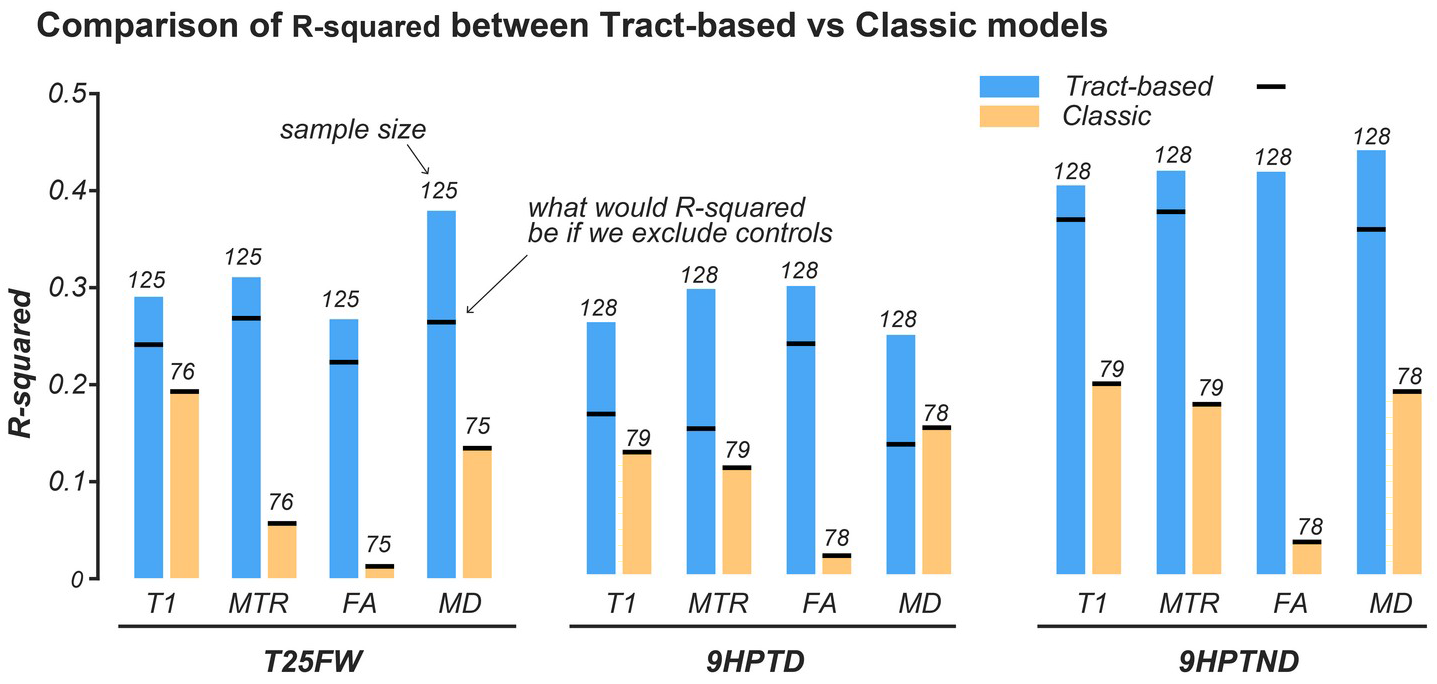
Comparison of explanatory power (R²) between tract-based and classical models for continuous disability outcomes (T25FW, 9HPT-D, 9HPT-ND), without demographic covariates. Bars are grouped by MRI metric (T1, MTR, FA, MD); blue bars indicate tract-based models and yellow bars indicate classical models. Statistical performance was assessed by ridge-penalized linear regression with L2 regularization (λ optimized by 10- fold cross-validation); Numbers above each bar indicate the sample size used in that analysis. Black horizontal lines indicate RZ values recomputed on sample-matched cohorts (healthy controls excluded to equate n between tract-based and classical models).

**Table 2.** Performance of ridge-penalized logistic regression models comparing tract-based and classical approaches for binary disability outcomes: EDSS > 3.0 (moderate impairment in at least one functional system) and MSPro ≥ 2 (elevated risk of disease progression), without demographic covariates. Models were fitted separately for each of four MRI metrics (T1, MTR, FA, MD) using ridge regression with L2 regularization; the regularization parameter (λ) was optimized by 10-fold cross-validation on training folds only. Model discrimination is reported as AUC (area under the ROC curve) with bootstrapped 95% confidence intervals (2000 resamples); higher AUC indicates better discrimination. Model fit is reported as AIC (Akaike Information Criterion); lower AIC indicates better model fit. Statistical significance of each model (unadjusted and Bonferroni-corrected across 20 comparisons) is provided in Supplementary Table 4. n denotes the number of participants with complete data for each model; values in parentheses indicate the number of participants in the positive class (EDSS > 3.0 or MSPro ≥ 2). Results including demographic covariates are provided in Supplementary Table 3.

| **Clinical Metric**<br>**(n=number of patients)** | **Tract-based**<br>**AUC (95% CI)** | **Classical-based**<br>**AUC (95% CI)** | **Tract-**<br>**based**<br>**AIC** | **Classical-based**<br>**AIC** |
| --- | --- | --- | --- | --- |
| EDSS-T1<br>(n=80) | 0.962<br>(0.911-1.000) | 0.850<br>(0.756-0.931) | 46.38 | 81.35 |
| EDSS-MTR<br>(n=80) | 0.906<br>(0.835-0.963) | 0.732<br>(0.613-0.841) | 63.99 | 100.95 |
| EDSS-FA<br>(n=79) | 0.897<br>(0.823-0.959) | 0.578<br>(0.440-0.707) | 67.22 | 111.91 |
| EDSS-MD<br>(n=79) | 0.902<br>(0.826-0.963) | 0.861<br>(0.771-0.936) | 66.40 | 81.16 |
| MSPro-T1<br>(n=79) | 0.893<br>(0.807-0.963) | 0.832<br>(0.739-0.919) | 67.09 | 85.70 |
| MSPro-MTR<br>(n=79) | 0.888<br>(0.810-0.953) | 0.660<br>(0.538-0.772) | 68.80 | 104.89 |
| MSPro-FA<br>(n=78) | 0.861<br>(0.772-0.931) | 0.568<br>(0.438-0.697) | 76.13 | 110.98 |
| MSPro-MD<br>(n=78) | 0.886<br>(0.803-0.952) | 0.709<br>(0.592-0.816) | 68.11 | 98.46 |

#### Baseline Associations of Lesion Burden with Disability

Correlations were obtained between the various lesion metrics and clinical disability scores. For ordinal clinical variables (EDSS, MSPro), Spearman’s rank correlation was used. For continuous timed variables (T25FW, 9HPT-D, 9HPT-ND), values were log-transformed prior to analysis to address positive skew, and Pearson correlation was then applied to the transformed values. To control for potential confounding effects, partial correlations were calculated using patient sex, age, and disease duration as covariates. For each lesion metric– clinical score pair, the correlation coefficient (r) is reported, and statistically significant correlations (p < 0.05) are indicated with an asterisk (*). Bonferroni correction was applied to control for multiple comparisons across correlation matrices (Supplementary Figure S1).

#### Multivariable Predictive Modelling: Tract-Based vs. Classical Approaches

Associations between MRI-derived metrics and clinical disability were modeled using ridge- penalized regression, comparing a Tract-Based model and a Classical-Based model. Binary outcomes representing clinically meaningful disability, EDSS > 3.0 (indicating moderate impairment in at least one functional system) and MSPro ≥ 2 (indicating elevated risk of disease progression), were analyzed using ridge-penalized logistic regression. Continuous outcomes (T25FW, 9HPT-D, 9HPT-ND) were analyzed with ridge-penalized linear regression. The regularization parameter (λ) was optimized using 10-fold cross-validation. Model performance was evaluated in the training cohort on the regularized models using AUC with bootstrapped 95% CIs and Akaike Information Criterion (AIC) for binary outcomes, and RZ (reported with and without healthy controls) for continuous outcomes. Covariates varied by the clinical outcome: age, sex, and disease duration for binary outcomes; age and sex for continuous outcomes (disease duration is excluded when healthy controls are included). Each analysis was performed both with and without demographic covariates. All analyses were conducted in MATLAB R2024b.^42^ For reference, identical multivariable analyses applied to whole brain lesion load metrics were also conducted. For the Tract-Based model, predictors were defined as tail percentiles from tract voxel distributions (10th percentile for MTR/FA; 90th percentile for T1/MD). These cut-offs reflect that reduced FA/MTR and elevated T1/MD signify demyelination and axonal disorganization. This approach avoided missing values in tracts without visible lesions on FLAIR scans, allowed inclusion of healthy controls, and captured the most abnormal voxels within each tract.

The Classical model used mean lesion values within the four predefined classical anatomical regions (see Fig. 3, top panel). Because lesion presence was required in all four regions, patients without complete lesion coverage were excluded. The resulting cohort size was reported for each analysis. Tail percentiles were not applied in the classical model, as lesion locations are undefined outside lesion masks. Because tail percentiles were drawn from the full tract distribution rather than restricted to FLAIR-visible lesions, NAWM microstructure contributes to the tract-based predictor in patients with sparse or absent lesions in a given cluster.

## Results

### Classical versus tract-based lesion load: Group-wise Comparison of Lesion Frameworks

#### Lesion Location

The overall lesion load in the Classical method represents the total lesion volume derived from whole-brain 3D FLAIR. Overall lesion load was lowest in RRMS, higher in PPMS, and more than doubled in SPMS as shown in Fig. 4a. Periventricular lesions accounted for the greatest share of this volume, followed by juxtacortical and deep-white-matter compartments, with infratentorial regions contributing the least. The tract-based metric, which includes only lesions intersecting predefined white-matter fiber bundles (Supplementary Table 2), captured 22% of the total lesion volume in RRMS, 37% in PPMS, and 27% in SPMS. Within these restricted tract-based masks, lesion burden rose progressively from RRMS through PPMS to SPMS, reproducing the phenotype-dependent escalation observed with the classical measure.

Tract-based lesion loads, especially the Lnorm, separate the three MS subtypes far more clearly than classical region-based metrics, while simultaneously highlighting the functional pathways most relevant to disease evolution (radar plots in Fig. 4b). Across all three tract-level measures, raw LN, LV and Lnorm, the distributions for RRMS, SPMS and PPMS are minimally or non-overlapping, mirroring the known escalation of disease burden. Lnorm, which adjusts for tract size and hemispheric side, produces the widest inter-quartile gaps. On the other hand, classical region-based metrics show inconsistent patterns, for instance the periventricular band, classical LN provides almost no separation between MS subtypes, whereas periventricular LV rises sharply, exaggerating group differences. The opposite pattern emerges in juxtacortical and deep-white-matter regions, where LN inflates differences and LV compresses them. In the analyzed tracts, lesion load was highest in association fibers, followed by projection-brainstem tracts, occipitoparietal bundles and cerebellar pathways.

#### Lesion Composition

Quantitative assessment of lesion composition using tract-based regions provided greater separation between MS subtypes than classical anatomical lesion mapping. As shown in Fig. 5, T1, MTR, FA, and MD values measured in lesions located within classical regions showed minimal differences between RRMS, PPMS, and SPMS groups. In contrast, tract-based lesion mapping showed subtype-specific trends in several white matter systems. The occipitoparietal tract demonstrated the most consistent separation across all tissue metrics when comparing MS subtypes. From RRMS to SPMS, we observed higher T1, lower MTR, higher MD, and lower FA. These directional changes across all four metrics reflect cumulative microstructural damage and provide a coherent tissue signature of progression within the occipitoparietal system. Other tract clusters demonstrated similar directionality but with varying magnitudes. In the cerebellar tract, FA and MTR values were greater in SPMS than PPMS, deviating from the trend seen in other regions.

Tract-based NAWM analysis showed a monotonic pattern across subtypes for all four metrics. T1 and MD values in occipitoparietal NAWM were progressively higher from control to RRMS, PPMS, and SPMS. MTR and FA values were correspondingly lower across these groups. These directional changes were observed in most tract clusters, including the projection-brainstem and association tracts. In contrast, cerebellar NAWM again deviated from this pattern in FA, with SPMS showing greater values than controls.

#### Baseline Associations of Lesion Burden with Disability

To establish a conventional clinical baseline for reference and future replication, we first report associations of lesion burden and disability using standard correlation analyses. At the global level, whole-brain lesion number did not reach statistical significance for any clinical outcome after Bonferroni correction. Whole-brain lesion volume showed significant correlation with EDSS (*r* = 0.51, adjusted *p* < 0.001), 9HPT-D (*r* = 0.40, adjusted *p* = 0.016), and 9HPT-ND (*r* = 0.60, adjusted *p* < 0.001), associate with either T25FW (*r* = 0.22, adjusted *p* = 1.00) or MSPro (*r* = 0.21, adjusted *p* = 1.00). Adjusted p-value is capped at 1.00 after correction.

We observed a similar pattern of results when splitting lesion burden into either tract-based or classical regions. The anatomical refinement did improve some specific correlation magnitude. For instance, tract-based lesion number within the cerebellar tracts significantly correlated with EDSS (*r* = 0.43) and deep white matter lesion number correlated with 9HPT-D (*r* = 0.51). A comprehensive table of all correlation results is provided in Supplementary Figure 1. In brief, lesion burden metrics showed moderate and outcome-dependent associations with disability: significant correlations were observed for EDSS and 9HPT, but not for T25FW or MSPro, for which no lesion burden metric reached significance after correction.

#### Multivariable Modeling of Lesion Location and Composition

To determine if integrating microstructural integrity and anatomical localization could better account for clinical variance, we built multivariable ridge-penalized regression models using four MRI metrics (T1, MTR, FA, and MD). A first goal of the multivariable predictive modeling was to evaluate whether tract-based frameworks outperform classical region-based approaches when predicting clinical disability in MS. Binary disability outcomes (Table 2) showed a clear advantage for the tract-based models. Across all MRI metrics, tract-based models achieved higher discrimination and better model fit than their classical counterparts for both EDSS and MSPro. For EDSS, tract-based AUCs ranged from 0.897 to 0.962 across MRI metrics (T1, MTR, FA, MD), compared with 0.578 to 0.861 for classical models. For MSPro, tract-based AUCs ranged from 0.861 to 0.893, compared with 0.568 to 0.832. Tract-based models also had consistently lower AIC values (e.g., EDSS-T1: 46.4 vs 81.4; MSPro-MTR: 68.8 vs 104.9).

Continuous outcomes (Fig. 6) demonstrated a similar pattern. Across all MRI metrics (T1, MTR, FA, MD) tract-based models explained more variance in clinical disability (T25FW, 9HPT-D, 9HPT-ND) than classical models. For example, the tract-based MD model explained 37.6% of the variance in the T25FW, notably outperforming the classical regional MD model (*R*^2^ = 0.135). Similarly, for 9HPT-D, the tract-based model provided the strongest predictive value (FA: *R*^2^ = 0.295). The strongest result for 9HPT-ND also came from tract-based models (MD: *R*^2^ = 0.434). The tract-based models utilized tail-based metrics, which allowed for the inclusion of healthy control subjects and thereby boosted the available sample size compared to the classical models. To verify that the higher explanatory power was not merely an artifact of this larger sample size, we also evaluated tract-based models using sample sets strictly matched (N-matched) to the classical cohorts, plotted as horizontal lines in Fig. 6. Even when constrained to equal sample sizes, the N-matched tract-based models consistently maintained an overall higher *R*^2^ values than the classical regional models.

In a supplementary analysis, we repeated the multivariable modeling with the inclusion of demographic covariates (Supplementary Table 3), we observed generally higher AUCs in nearly all binary models and in *R*^2^ across all continuous models after the inclusion of demographic covariates (Supplementary Figure S2). Consistent with previous results, the tract-based results consistently outperformed the classic model. Following Bonferroni correction (Supplementary Table 4), most tract-based models (19 out of 20) remained statistically significant for all clinical outcomes except for MSPro-MTR where the adjusted p-value was 0.0596. In contrast, only 4 of the 20 classical regional models survived correction and all metrics failed for MSPro, T25FW and 9HPT-D. Although a few classical models did come very close (9HPTD-MD, adjusted *p* = 0.0528; 9HPTND-T1, adjusted *p* = 0.0541; 9HPTND- MD, adjusted *p* = 0.0549). For reference, multivariable analyses applied to whole brain lesion load metrics are provided in Supplementary Table 5.

## Discussion

### Study recap

This study aimed to improve the clinical relevance of lesion analysis in MS by combining tract-based spatial filtering with quantitative microstructural MRI metrics.

We hypothesized that filtering lesions through clusters of functionally meaningful white matter tracts rather than broad anatomical compartments would allow for more accurate identification of clinically relevant damage. The premise is intuitive: lesions that disrupt specific tracts or their intersections likely carry greater functional weight than those in other regions. Such filtering provides a biologically grounded and reproducible framework for lesion inclusion and exclusion, enabling more meaningful analysis of tissue properties within these tracts.

The clinico-radiological paradox in MS persists because not all lesions are the same. While lesion location has traditionally been used to explain disability,^43,44^ our findings confirm that lesion position alone, even when filtered through functionally relevant white matter tracts, offers limited explanatory power. This is expected: lesion number, volume, or normalized volume may reflect spatial patterns of disease but cannot differentiate between biologically distinct lesion types.

Consistent with this, Vellinga et al.^45^ showed that in a large voxelwise analysis of MS patients, correlations between lesion probability and disability were largely confined to periventricular regions and were strongly influenced by total lesion load (LL), which itself correlated with lesion probability across the white matter. The authors suggested that these apparent location– disability relationships may therefore be driven by overall lesion burden rather than truly localized effects. This highlights an important limitation of conventional T2-based approaches: although spatial mapping refines simple lesion counts, it remains constrained by the biological non-specificity of treating all lesions as equivalent. Our framework addresses these limitations by jointly incorporating anatomical specificity (restricting analysis to functionally relevant tracts) and microstructural composition through qMRI tail metrics.

### Tract-Based Filtering and the Value of Lesion Location

MS lesions are both spatially and biologically heterogeneous, varying across patients and within the same individual. A single patient may harbor lesions at distinct anatomical locations that differ in their functional importance, while at the same time, lesions at different disease stages (e.g., RRMS vs. SPMS) differ in composition, chronicity, and impact on disability.^46^ This dual heterogeneity, of where lesions occur and what they contain, lies at the core of the paradox originally described by Barkhof, in which conventional MRI lesion load fails to reflect clinical severity.^9^

Hence, capturing MS disease severity and progression with quantitative MRI hinges on two complementary dimensions: lesion topography (precise spatial distribution) and lesion microstructure (intrinsic pathology such as demyelination, axonal transection, and gliosis). Nonetheless, anatomically defined ROIs, like the periventricular compartment, treat every lesion that contacts the ependymal surface as equivalent, regardless of its microstructural damage or functional connectivity. Because the periventricular mask can extend far from the ventricular lining whenever plaques merely “touch” the ROI, confluent lesions in SPMS disproportionately inflate total burden and obscure stage-specific changes. Likewise, classical anatomical masks potentially include lesions irrelevant to key neural circuits while excluding strategically disruptive foci.

By restricting analysis to lesions within well-characterized, functionally important white-matter tracts, tract-based filtering reduces this noise and sharpens specificity. As illustrated in Fig. 4a, this approach isolates the spatial zones where lesion microstructure mirrors the connections/disconnections inside an ROI. Moreover, this framework now enables analysis of NAWM within restricted dimensions defined by each tract, thereby reflecting underlying biological and molecular heterogeneity.

This principle aligns with the work of Harrison et al.,^24^ who demonstrated that tract-specific DTI and MTR metrics, restricted to the optic radiation, corticospinal tract, and corpus callosum, correlated better with EDSS than whole-brain lesion load. Importantly, these relationships were not uniform across disease stages: significant correlations between tract-specific metrics and disability were observed in RRMS but were absent in SPMS, suggesting a stage-dependent dissociation between structural damage and clinical expression. These findings support the notion that the relationship between white matter pathology and disability is both anatomically specific and modulated by disease progression. The importance of anatomical precision is further illustrated by Madsen et al.,^21^ who used ultra-high field 7T MRI to demonstrate that the presence of cortical lesions within the somatotopically defined primary sensorimotor hand area (SM1-HAND) is associated with contralateral motor and sensory impairment, as well as altered corticospinal conduction and excitability. Importantly, these relationships remained significant after accounting for other MRI-derived measures of pathway damage, and were spatially specific, as lesions in control regions showed no such effects. While whole-brain cortical lesion load was also associated with global disability, the regional analysis provided greater domain-specific sensitivity.

Extending this further, Kerbrat et al.^47^ mapped lesions along the corticospinal tract from the motor cortex to the cervical cord in 290 MS patients across multiple MS subtypes, demonstrating that baseline spinal cord CST lesion volume was the only variable independently associated with risk of EDSS progression at 2 years. Similarly, Kim et al. ^22^ proposed a tract density index weighting lesions by the fiber density of intersected tracts, which correlated with EDSS (r = 0.30). While our framework differs methodologically; encoding microstructural composition rather than connectivity weighting, both approaches converge on the same principle: the network relevance of lesion location matters more than raw lesion burden, and our AIC-confirmed model advantage provides independent multivariable support for this conclusion.

### Integrating Tissue Composition Enhances Clinical Associations

Incorporating quantitative MRI modalities (T1, MTR, and diffusion) into the tract-based framework, we sought to characterize underlying lesion pathology and evaluate their predictive value for clinical disability. While previous studies reported modest correlations between mean T1 values of whole-brain white matter lesions and EDSS,^48^ or between MTR of cortical lesions and EDSS,^49^ these approaches treated all lesions equally. In contrast, tract-based analysis minimizes contributions from silent regions and amplifies signals from lesions in functionally critical white matter pathways.

Harper et al.^48^ reported no significant cross-sectional association between median T1 values and clinical disability in early RRMS, attributing this null finding in part to whole-lesion T1 pooled across broad anatomical compartments. Our tract-based T1 tail metric showed meaningful associations with motor outcomes across subtypes, including binary EDSS and MSPro classification. The discrepancy is methodologically informative: aggregating T1 across anatomically heterogeneous lesion pools dilutes the signal from pathologically and functionally consequential lesions. Restricting sampling to tract-defined tissue substantially recovers that signal.

A parallel limitation is evident in the MTR literature. Amann et al.^49^ found that MTR of cortical lesions showed only a trend toward EDSS correlation, and critically no MTR parameter predicted T25FW or 9HPT in multivariable analysis. Our study demonstrates significant associations between tract-based MTR and all three motor outcomes, including gait (T25FW) and manual dexterity (9HPT-D and 9HPT-ND). This reversal of a previously limited and domain-specific association underscores that the anatomical framework in which MTR is measured, not the metric itself, enhances its clinical sensitivity. Restricting MTR sampling to white matter tracts with known functional relevance appears to be a key contributing factor, enabling its translation to motor disability prediction.

The multivariable models used tract-specific tail percentile metrics that simultaneously encode tissue composition and anatomical location. The consistent performance advantage of tract-based over classical models was sustained across all four MRI metrics, both binary outcomes, and all three continuous functional measures. The lower AIC values confirm that this advantage did not arise from overfitting or added model complexity. This suggests that anchoring MRI measurements to coherent fiber systems yields a more specific representation of clinically relevant white matter damage than pooling lesion statistics across anatomically defined but functionally heterogeneous regions.

Diffusion-based metrics, in particular, illustrate the value of anchoring MRI measurements to coherent fiber systems. FA and MD reflect directional fiber coherence and water diffusivity, properties that are biologically meaningful along the principal axes of white matter tracts but become harder to interpret when measured across heterogeneous lesion pools spanning multiple anatomical regions. Measured within tract-defined boundaries, diffusion metrics showed the largest performance gap between tract-based and classical models. For FA and MD specifically, the performance advantage over classical ROI-based diffusion metrics suggests the benefit is driven by the spatial restriction to functionally relevant tracts rather than by any specific diffusion-sensitive contrast. The tract-based results for T25FW explained a substantial amount of variance, suggesting that spatially localized white matter damage along motor-relevant tracts is a sensitive predictor of gait performance.

Beyond overt lesion microstructure, our framework captures NAWM properties within tract- defined boundaries, consistent with the paradigm established by Ravano et al.,^15^ who demonstrated that tract-wise T1 deviations in both lesions and NAWM are associated with current disability and predict future disability progression in early MS. Notably, microstructural alterations in specific pathways, particularly infratentorial tracts, were more strongly linked to disability evolution than cross-sectional disability, highlighting the prognostic relevance of covert tissue damage within functionally defined networks. Our approach extends this tract-restricted sampling strategy to a full MS spectrum cohort using four qMRI metrics, and the consistent performance advantage of tract-based over classical models across clinical outcomes reflects the same underlying principle: microstructural damage in functionally critical white matter, often invisible to conventional MRI, carries clinical information precisely because its anatomical context is preserved.

Tract-specific DTI metrics had shown early promise, but prior work remained limited by univariate analyses and single clinical outcomes.^24^ Lapucci et al.^50^ specifically demonstrated disconnectome volume as the sole independent EDSS predictor in a multivariable model, providing direct empirical support for the primacy of network-referenced lesion metrics. We have shown here that anchoring T1, MTR, FA, and MD sampling to functionally relevant white matter tracts, and encoding both spatial and microstructural lesion attributes simultaneously in a multivariable framework, translates these individually weak signals into robust, AIC- confirmed predictors of motor disability across the full MS phenotype spectrum. Furthermore, LASSO-based DTI analyses predicting four-year motor outcomes (T25FW, 9HPT) from baseline tract-specific FA provide longitudinal support for the structure–function relationships we report cross-sectionally, suggesting that the microstructural associations identified here may have meaningful prognostic value.^51^ Disability accumulation in MS reflects both relapse-associated worsening and progression independent of relapse activity (PIRA), yet clinical relapses capture only the overt fraction of focal inflammation and carry no information about where damage falls relative to function. By referencing lesions to functionally critical tracts, our framework offers a spatially and functionally resolved read-out of focal injury that may reflect this biology more faithfully than relapse-based phenotyping alone, but testing this will require longitudinal study, ideally combined with markers of chronic active inflammation (paramagnetic rim or slowly expanding lesions) and neuroaxonal loss (atrophy, neurofilament light).

Our findings indicate that the tract-based framework provides a reliable substrate for detecting structure-function associations regardless of the specific MRI metrics used. It is important to note, however, that exact p-values carry limited interpretive value in isolation without real repeated replication studies from other groups. The exact results reported here reflect only one dataset, one preprocessing pipeline, and one set of modelling choices. Clinical neuroimaging research carries inherent uncertainty at every stage, and these findings should be treated as a starting point rather than a definitive benchmark. In that regard, replication studies can benefit the field as much as and if not more than novel studies. The longer-term goal is a shift toward individualized radiological reporting, where clinically available imaging data is interpreted in a way that is meaningful and actionable at the level of the individual patient. The scope, limitations, and directions for replication are discussed further in the following section.

### Methodological Considerations and Future Directions

While our findings highlight the clinical relevance of tract-informed lesion and NAWM metrics, several methodological limitations merit consideration. From a reproducibility standpoint, both MP2RAGE-based T1 mapping and MTR pose challenges for multi-center implementation. MP2RAGE, though offering superior tissue contrast and quantitative stability, is not yet widely available and requires longer acquisitions. Likewise, MTR measures show marked inter-vendor variability even under standardized conditions, reflecting incomplete harmonization of acquisition and processing workflow.^52^ These limitations constrain cross-site comparability of quantitative T1 and MTR metrics and limit their large-scale clinical adoption. Ongoing efforts toward vendor-neutral sequences and transparent workflows are essential to improve reproducibility and facilitate broader use in MS research.^52^ A more practical approach is to use DTI metrics (FA and MD) to quantify lesion composition. DTI is likely the preferred method, as it provides both tract-based lesion localization and compositional information through FA and MD measures.

Future work combining tract-specific microstructural markers with lesion disconnectome metrics, quantifying the total volume of white matter tracts intersected by lesions,^50^ could further improve disability prediction, as both approaches have independently shown superiority over anatomical ROI methods.^24^ Similarly, longitudinal replication of the tract-wise microstructural associations identified here, particularly in early or pre-symptomatic MS stages, would establish whether these cross-sectional relationships reflect static damage or ongoing microstructural evolution.^15,16^

Also, we would like to note that several analytical thresholds and parameter choices in the present study were selected on pragmatic rather than data-driven grounds, and we acknowledge these as limitations in the current work. The percentile cutoff used to define the tract-based tail metric, the binarization thresholds for EDSS and MSPro, and the choice of predictor sets and tract groupings were all fixed a priori based on clinical convention or interpretability rather than optimized within the data. In principle, each of these could be tuned as a free parameter; in practice, doing so across five outcomes, four imaging metrics, and multiple model families would substantially increase the risk of overfitting and complicate interpretation. We therefore view these choices as an analytical constraint for the current work that imposed a consistent, pre-specified structure across all analyses, at the cost of some loss of sensitivity for any individual comparison. Validation in independent cohorts with prospectively defined thresholds will be more than necessary in future studies. Furthermore, while ridge regression was applied specifically to reduce the risk of over-parameterisation by shrinking coefficients, we acknowledge that some optimism in the reported AUC estimates may remain, particularly given the modest sample size relative to the number of predictors. Calibration slopes were not formally assessed in the current study, and their evaluation, alongside external validation in independent cohorts, remains an important step before these models could be considered for clinical translation.

More specifically on the point regarding the choice of predictors, the decision of how to define tract groupings was not straightforward. We considered analyzing individual tracts as separate predictors, but ultimately grouped them into four functional clusters using the default atlas parcellation in DSIStudio. Treating each of the many individual tracts as a standalone feature would effectively transform this study into a data-driven feature selection problem and one that is better suited to a dedicated machine learning study with a substantially larger dataset. The current sample size does not provide sufficient statistical power for reliable tract-specific inference in a multivariable framework, and the risk of spurious associations would be considerable. That said, Fig. 5 offers a preliminary indication that the four groups do not contribute equally, and a reasonable a priori hypothesis is that sensorimotor tracts (such as those in occipitoparietal or projection-brainstem region) would contribute disproportionately to predictions of gait and upper limb function. Future work with larger cohorts should test this explicitly and explore more granular or data-driven tract parcellations beyond the default atlas groupings. Similarly, we evaluated each MRI metric independently rather than combining modalities, for example using T1 alongside FA in a single model. Such combinations could in principle capture complementary tissue properties, but the number of possible multi-metric combinations grows quickly, the resulting models become harder to interpret, and in routine clinical practice it is unlikely that a patient will have all modalities available simultaneously. Systematic multiparametric integration is therefore left for future work.

More broadly, while our tract-based framework enables refined tissue sampling, future work should further tailor tract filtering to specific clinical endpoints rather than applying universal lesion mask across all outcomes. The inclusion of spinal cord atrophy measures and advanced imaging modalities such as quantitative susceptibility mapping could improve sensitivity to iron dynamics and identifying “smoldering” or chronic active lesions, particularly in progressive MS.^53,54^ Longitudinal studies will also be critical to validate whether these tract-specific markers predict clinical decline over time. Follow-up imaging will allow us to assess whether early qMRI abnormalities evolve in parallel with symptom progression, helping define actionable thresholds. Finally, methodological advances in cerebellar tract segmentation and motion correction will likely enhance correlation detection in regions currently affected by signal instability or tracking artifacts.^55^

## Conclusion

This study demonstrated that combining lesion location with microstructural composition through tract-based qMRI mapping offers a clinically meaningful advancement in characterizing MS-related disability. The integration of spatial and biological lesion attributes translates into clinically informative associations between imaging markers and functional outcomes beyond what conventional lesion measures provide. As quantitative MRI becomes standardized and computational modelling more accessible, this framework supports the development of imaging-driven stratification tools for research and future clinical translation. Ultimately, we believe this approach transforms lesion analysis from a descriptive exercise into a functionally contextualized imaging marker, bridging radiological findings with clinical outcomes.

## Data Availability

Data produced in the present study are available upon reasonable request to the authors

## Acknowledgements

This study was supported by the Core Technology Platforms (CTP), Biomedical Imaging Core at New York University Abu Dhabi (NYUAD). We also acknowledge the Computational Research Center at NYUAD for providing supercomputing resources.

## Data Availability

The clinical and imaging data underlying this article cannot be shared publicly due to the inclusion of potentially identifying patient information; de-identified data supporting the analyses in this manuscript are available from the corresponding author on reasonable request, subject to approval by the Local Ethics Committee. The MATLAB analysis code is available at https://github.com/putiw/MS100Public.git.

---

**Supplementary Figure S1.**
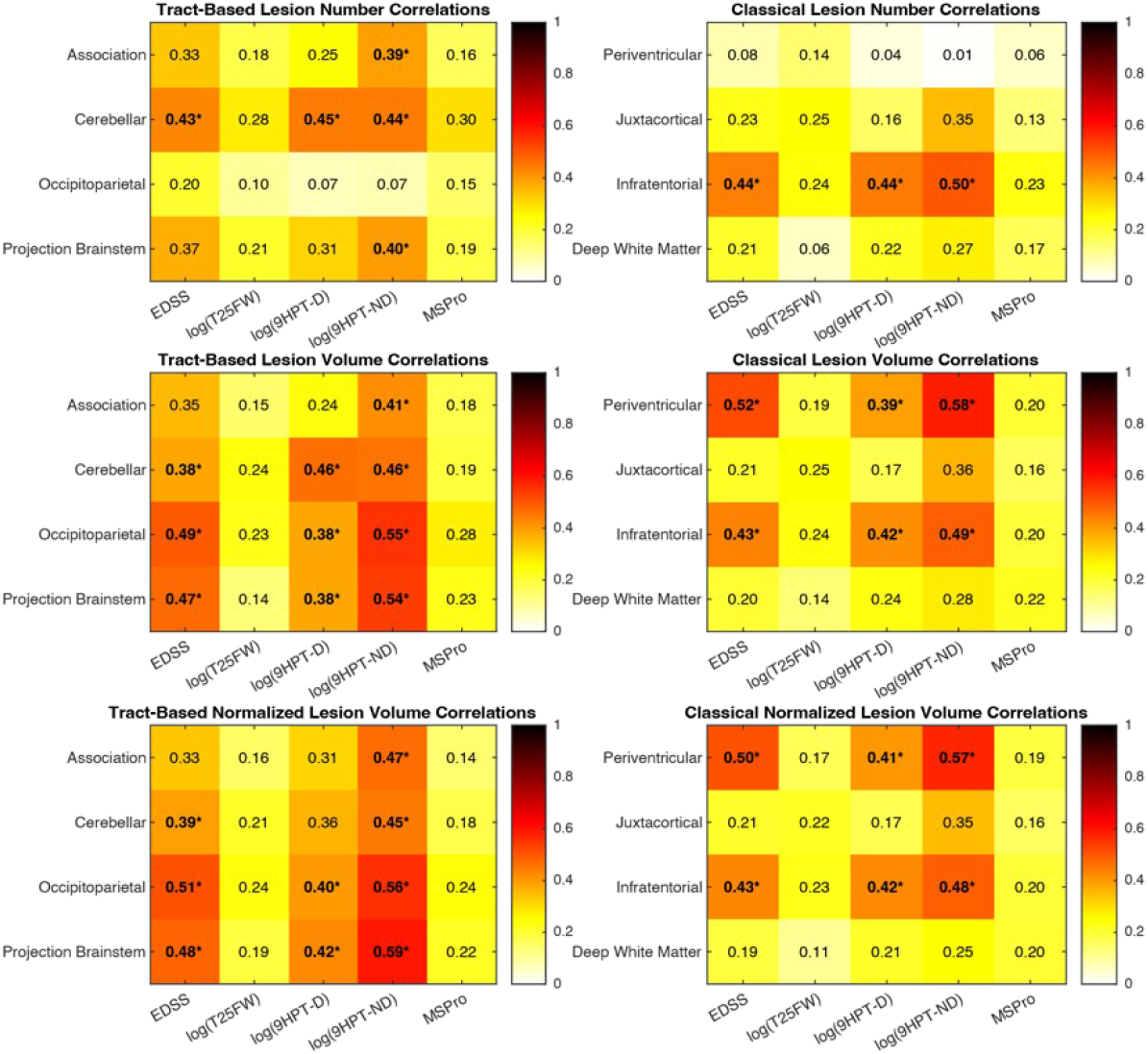
Partial correlations between lesion location metrics (lesion number [LN], lesion volume [LV], normalized lesion volume [Lnom]) and clinical disability scores, across tract-based and classical lesion frameworks. Partial correlations were computed controlling for patient age, sex, and disease duration as covariates; healthy controls were excluded from correlation analyses. For ordinal outcomes (EDSS, MSPro), Spearman’s rank correlation coefficient (ρ) was used. For continuous timed outcomes (T25FW, 9HPT-D, 9HPT-ND), values were log-transformed prior to analysis to address positive skew, and Pearson’s correlation coefficient (r) was applied to the transformed values. P-values were adjusted for multiple comparisons across all tests using Bonferroni correction; statistically significant associations (p_adj < 0.05 after correction) are annotated with (*). Each correlation was computed across MS patients (N = 89).

**Supplementary Figure S2.**
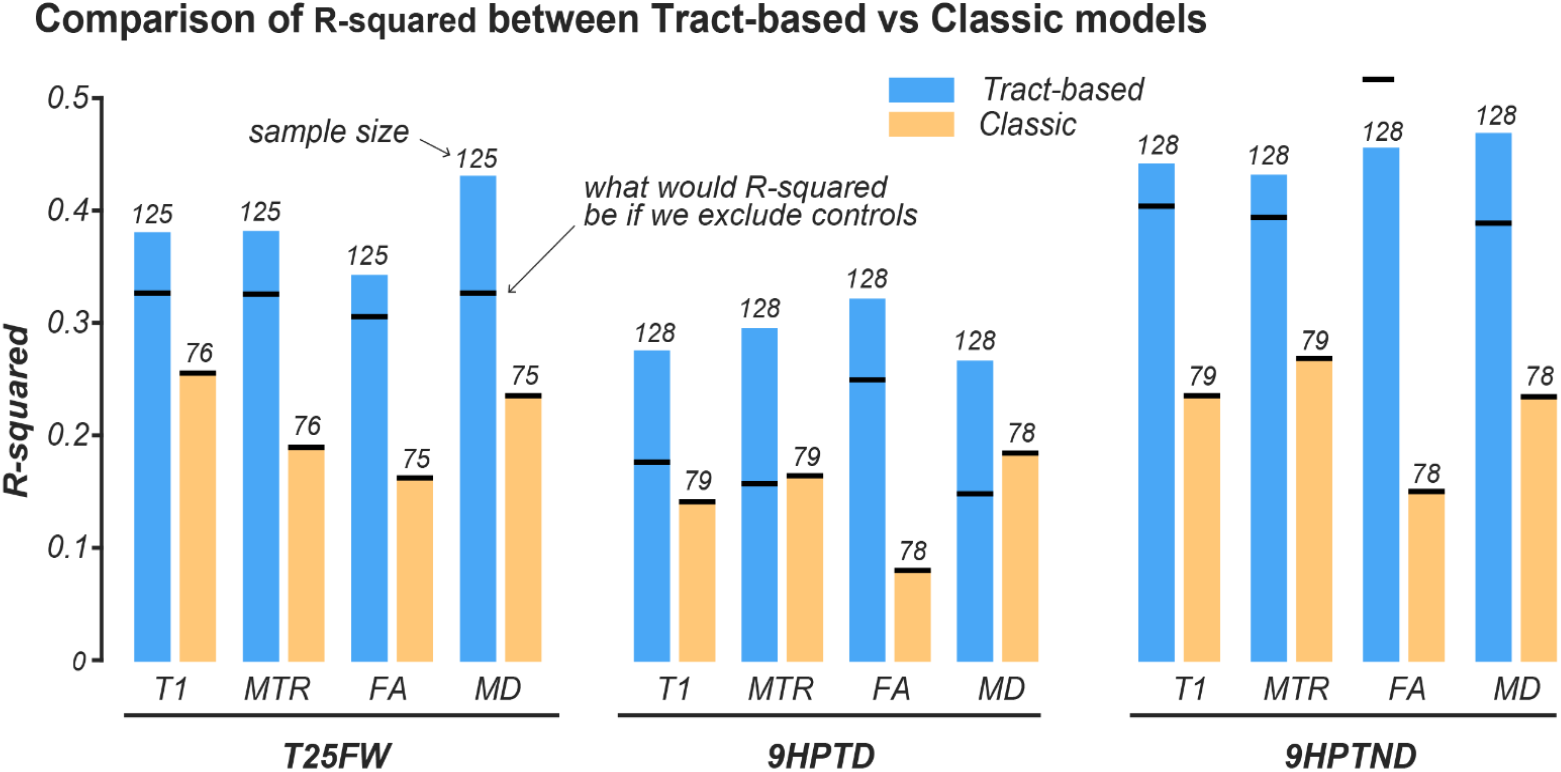
Comparison of explanatory power (R²) between tract-based and classical models for continuous disability outcomes (T25FW, 9HPT-D, 9HPT-ND), including demographic covariates. Because healthy controls are included in the continuous-outcome analyses and do not have a disease duration, demographic covariates were limited to age and sex; disease duration was included only in the binary-outcome models (EDSS, MSPro; MS patients only; see Supplementary Table 3). Bars are grouped by MRI metric (T1, MTR, FA, MD); blue bars indicate tract-based models and yellow bars indicate classical models. Statistical performance was assessed by ridge-penalized linear regression with L2 regularization (λ optimized by 10-fold cross-validation). Numbers above each bar indicate the sample size used. Black horizontal lines indicate RZ values recomputed on sample-matched cohorts (controls excluded). Bonferroni- corrected p-values are reported in Supplementary Table 4.

**Supplementary Table 1.** Comparative overview of the five disability assessment tools employed in this study: EDSS, T25FW, 9HPT (dominant, 9HPT-D, and non-dominant hands, 9HPT-ND), and MSPro. Each scale is characterized along three dimensions: (1) functional domains assessed, (2) task description, and (3) scoring system.

| Scale | Expanded Disability Status Scale (EDSS) | Timed 25-Foot Walk (T25FW) | Nine-Hole Peg Test (9HPT) | MSPRO |
| --- | --- | --- | --- | --- |
| **Functional Domains** | **Pyramidal** (muscle weakness or difficulty moving limbs)<br>**Cerebellar** (coordination)<br>**Brainstem** (speech and eye movement)<br>**Sensory** (numbness or loss of sensations)<br>**Bowel** and bladder function<br>**Visual** function<br>**Cerebral** or mental function<br>**Ambulation** (walking and use of aids) | **Ambulation**<br>**Lower limb** strength<br>**Balance** and motor control | **Fine motor** coordination<br>**Finger dexterity**<br>**Visual-motor** integration<br>**Bilateral upper limb** function (dominant and non-dominant hands)<br>**Independence** in activities of daily living (ADLs) | **Disease activity** (relapse frequency)<br>**Symptoms:** Motor, visual, coordination/balance, speech, sensory, bladder/bowel<br>**Cognitive** and fatigue-related functions<br>**Mobility**, self-care, occupational functioning, hobbies, and social engagement |
| **Type of Tasks** | **Motor tasks:** Evaluating muscle strength and coordination<br>**Sensory tests:** Assessing response to touch, pain, and vibration<br>**Visual assessments:** Testing visual acuity and field<br>**Gait analysis:** Measuring walking distance and stability | The individual is instructed to walk 25 feet as quickly and safely as possible. The time taken to complete the walk is recorded. | Pick up nine pegs one at a time and place them into nine holes on a pegboard as quickly as possible. Remove them one at a time and return them to the container. Each hand is tested separately. The task is completed under standardized conditions. | Symptom History (last 6 months)<br>Impact Assessment impact severity on areas of functioning. |
| **Scoring System** | 0.0–3.0: No to moderate disability but is fully ambulatory<br>3.5–5.5: Increasing difficulty walking but still ambulatory without aid for 300+ meters<br>6.0–7.5: Ambulation severely restricted; requires assistance (e.g., cane, walker, or wheelchair)<br>8.0–9.5: Essentially restricted to bed or chair; retains some self-care functions<br>10.0: Death due to MS | Healthy Controls: Median T25FW time is approximately 3.7 seconds (range: 2.8–5.2 seconds)<br>20% or greater increase in T25FW time is considered a clinically meaningful change, indicating potential disease progression | Normative Data: Norms vary based on age and sex. Cut-off values:<br>>18 seconds is generally considered abnormal for the dominant hand.<br><0.5 pegs/s may indicate high risk for activity limitations.<br>33.3 seconds (corresponding to ~0.27 pegs/s) separates mild from more severe impairment | Section A: Relapse and Recovery (max 14 points)<br>Section B: Symptoms (max 91 points)<br>Section C: Impacts (max 14 points)<br>Total Score = Sum of all above sections<br>Interpretation Output:<br>1: (<46.3): Unlikely progression<br>2: (46.3–57.8): Possible progression<br>3: (>57.8): Likely progression |

**Supplementary Table 2.** Assignment of 24 canonical white matter tracts to four functional clusters (occipitoparietal, association, projection-brainstem, cerebellar) used in tract-based lesion analyses. Tract parcellation was performed using DSI Studio’s ICBM-152 template-based deterministic tracking.

| **Occipitoparietal** | **Association** | **Projection-Brainstem** | **Cerebellar** |
| --- | --- | --- | --- |
| Projection Basal Ganglia: Optic Radiation L&R | Arcuate Fasciculus L&R | Corticospinal Tract L&R | Inferior Cerebellar Peduncle L&R |
| Association Vertical Occipital Fasciculus L&R | Cingulum Frontal Parahippocampal L&R | Corticobulbar Tract L&R | Middle Cerebellar Peduncle |
| Corpus Callosum: Forceps Major | Cingulum Parahippocampal L&R | Corticopontine Tract-Frontal L&R | Superior Cerebellar Peduncle |
|  | Uncinate Fasciculus L&R | Corticopontine Tract-Parietal L&R | Cerebellum White Matter L&R |
|  | Superior Longitudinal Fasciculus 2 L&R | Corticopontine Tract-Occipital L&R | Vermis |
|  | Superior Longitudinal Fasciculus 3 L&R | Medial Lemniscus L&R |  |
|  | Corpus Callosum: Forceps Minor | Reticular Tract L&R |  |
|  |  | Corpus Callosum: Body |  |
|  |  | Corpus Callosum: Tapetum |  |

**Supplementary Table 3.** Performance of ridge-penalized logistic regression models comparing tract-based and classical approaches for binary disability outcomes (EDSS > 3.0 and MSPro ≥ 2), including demographic covariates (age, sex, and for MS patients only: disease duration). Statistical methods are identical to those described for Table 2. The regularization parameter (λ) was optimized by 10-fold cross-validation on training folds only. Model discrimination: AUC (bootstrapped 95% CI, 2000 resamples). Model fit: AIC. n denotes participants with complete data; values in parentheses indicate the positive class count. Bonferroni-corrected p-values for all models are reported in Supplementary Table 4.

| **Clinical Metric**<br>**(n=number of subjects)** | **Tract-Based AUC**<br>**(95% CI)** | **Classical-Based**<br>**AUC (95% CI)** | **Tract-**<br>**Based AIC** | **Classical-**<br>**Based AIC** |
| --- | --- | --- | --- | --- |
| **EDSS-T1**<br>**(n=80)** | 0.956<br>(0.904-0.999) | 0.885<br>(0.803-0.957) | 47.83 | 79.87 |
| **EDSS-MTR**<br>**(n=80)** | 0.919<br>(0.853-0.971) | 0.871<br>(0.787-0.941) | 66.31 | 81.93 |
| **EDSS-FA**<br>**(n=79)** | 0.935<br>(0.876-0.978) | 0.786<br>(0.679-0.881) | 60.81 | 95.93 |
| **EDSS-MD**<br>**(n=79)** | 0.923<br>(0.859-0.973) | 0.897<br>(0.822-0.959) | 67.11 | 76.19 |
| **MSPro-T1**<br>**(n=79)** | 0.989<br>(0.970-1.000) | 0.934<br>(0.877-0.983) | 31.62 | 65.18 |
| **MSPro-MTR**<br>**(n=79)** | 0.979<br>(0.950-0.999) | 0.929<br>(0.866-0.978) | 38.20 | 64.46 |
| **MSPro-FA**<br>**(n=78)** | 0.966<br>(0.926-0.993) | 0.922<br>(0.852-0.976) | 45.71 | 65.95 |
| **MSPro-MD**<br>**(n=78)** | 0.965<br>(0.917-0.996) | 0.926<br>(0.859-0.978) | 47.06 | 64.62 |

**Supplementary Table 4.** Unadjusted and Bonferroni-corrected p-values for tract-based and classical ridge-penalized regression models across all five clinical outcomes (EDSS, MSPro, T25FW, 9HPT-D, 9HPT-ND) and four MRI metrics (T1, MTR, FA, MD), yielding 20 model comparisons per approach. Binary outcomes (EDSS, MSPro) were modeled with ridge-penalized logistic regression; continuous outcomes (T25FW, 9HPT-D, 9HPT-ND) were modeled with ridge-penalized linear regression. The regularization parameter (λ) was optimized by 10-fold cross-validation. P-values were derived from the cross-validated model fit; Bonferroni correction was applied across the 20 model comparisons (α_adj = 0.05/40 = 0.00125). N denotes total participants; values in parentheses indicate the positive-class count for binary outcomes. p_adj values capped at 1.000 after correction.

| Outcome | MRI Metric | Model | N | p | p_adj |
| --- | --- | --- | --- | --- | --- |
| EDSS | T1 | Tract-based | 80 (39) | **0.0002** | **0.0074** |
|  |  | Classic | 80 (39) | **0.0003** | **0.0122** |
|  | MTR | Tract-based | 80 (39) | **< 0.0001** | **0.0012** |
|  |  | Classic | 80 (39) | **0.0005** | **0.0207** |
|  | FA | Tract-based | 79 (39) | **< 0.0001** | **0.0019** |
|  |  | Classic | 79 (39) | 0.1805 | 1.0000 |
|  | MD | Tract-based | 79 (39) | **0.0001** | **0.0037** |
|  |  | Classic | 79 (39) | **0.0008** | **0.0301** |
| MSPro | T1 | Tract-based | 79 (39) | **0.0010** | **0.0402** |
|  |  | Classic | 79 (39) | 0.0339 | 1.0000 |
|  | MTR | Tract-based | 79 (39) | **0.0015** | **0.0596** |
|  |  | Classic | 79 (39) | 0.0233 | 0.9339 |
|  | FA | Tract-based | 78 (39) | **0.0009** | **0.0363** |
|  |  | Classic | 78 (39) | 0.0653 | 1.0000 |
|  | MD | Tract-based | 78 (39) | **0.0008** | **0.0331** |
|  |  | Classic | 78 (39) | 0.0405 | 1.0000 |
| T25FW | T1 | Tract-based | 125 | **< 0.0001** | **< 0.0001** |
|  |  | Classic | 76 | 0.0024 | 0.0947 |
|  | MTR | Tract-based | 125 | **< 0.0001** | **0.0000** |
|  |  | Classic | 76 | 0.0784 | 1.0000 |
|  | FA | Tract-based | 125 | **< 0.0001** | **< 0.0001** |
|  |  | Classic | 75 | 0.4016 | 1.0000 |
|  | MD | Tract-based | 125 | **< 0.0001** | **< 0.0001** |
|  |  | Classic | 75 | 0.0070 | 0.2786 |
| 9HPT-D | T1 | Tract-based | 128 | **< 0.0001** | **< 0.0001** |
|  |  | Classic | 79 | 0.0110 | 0.4401 |
|  | MTR | Tract-based | 128 | **< 0.0001** | **< 0.0001** |
|  |  | Classic | 79 | 0.0034 | 0.1349 |
|  | FA | Tract-based | 128 | **< 0.0001** | **< 0.0001** |
|  |  | Classic | 78 | 0.2164 | 1.0000 |
|  | MD | Tract-based | 128 | **< 0.0001** | **< 0.0001** |
|  |  | Classic | 78 | **0.0013** | **0.0528** |
| 9HPT-ND | T1 | Tract-based | 128 | < 0.0001 | < 0.0001 |
|  |  | Classic | 79 | 0.0014 | 0.0541 |
|  | MTR | Tract-based | 128 | < 0.0001 | < 0.0001 |
|  |  | Classic | 79 | 0.0002 | 0.0093 |
|  | FA | Tract-based | 128 | < 0.0001 | < 0.0001 |
|  |  | Classic | 78 | 0.1113 | 1.0000 |
|  | MD | Tract-based | 128 | < 0.0001 | < 0.0001 |
|  |  | Classic | 78 | 0.0014 | 0.0549 |

*p*: unadjusted *p*-value (Wald test). *p<sub>adj</sub>*: Bonferroni corrected *p*-value, applied per family of 40 tests. EDSS: Expanded Disability Status Scale; MSPro: MS Progression; T25FW: Timed 25-Foot Walk; 9HPT-D/ND: 9-Hole Peg Test dominant/non-dominant hand.

**Supplementary Table 5.** Model performance (AUC [95% CI] and AIC for binary outcomes; RZ and AIC for continuous outcomes) of ridge-penalized regression models using lesion count and volume metrics (whole-brain LN, whole-brain LV, tract-based LN/LV/Lnom,, and classical LN/LV/Lnom) as predictors, without demographic covariates. These analyses serve as a reference baseline for comparison with the microstructural composition models in Table 2 and Supplementary Table 3. Statistical methods are identical to those described for Table 2: ridge-penalized logistic regression (binary) or linear regression (continuous) with λ optimized by 10-fold cross-validation; AUC with bootstrapped 95% CI (2000 resamples).

| Outcome | Metric | Model | N | AUC (95% CI) | AIC |
| --- | --- | --- | --- | --- | --- |
| EDSS | LN | Whole Brain | 89 (41) | 0.673 [0.556, 0.787] | 122.45 |
|  | LV | Whole Brain | 89 (41) | 0.828 [0.734, 0.908] | 97.09 |
|  | LN | Tract-based | 89 (41) | 0.840 [0.748, 0.915] | 101.82 |
|  | LN | Classic | 89 (41) | 0.693 [0.579, 0.806] | 116.75 |
|  | LV | Tract-based | 89 (41) | 0.872 [0.786, 0.942] | 84.05 |
|  | LV | Classic | 89 (41) | 0.833 [0.736, 0.914] | 91.76 |
|  | Lnorm | Tract-based | 89 (41) | 0.864 [0.781, 0.933] | 87.28 |
|  | Lnorm | Classic | 89 (41) | 0.823 [0.723, 0.908] | 93.18 |
| MSPro | LN | Whole Brain | 88 (42) | 0.619 [0.494, 0.734] | 123.11 |
|  | LV | Whole Brain | 88 (42) | 0.693 [0.579, 0.803] | 117.97 |
|  | LN | Tract-based | 88 (42) | 0.704 [0.587, 0.808] | 115.75 |
|  | LN | Classic | 88 (42) | 0.639 [0.524, 0.754] | 120.15 |
|  | LV | Tract-based | 88 (42) | 0.762 [0.655, 0.855] | 103.76 |
|  | LV | Classic | 88 (42) | 0.707 [0.592, 0.815] | 116.43 |
|  | Lnorm | Tract-based | 88 (42) | 0.739 [0.628, 0.841] | 111.75 |
|  | Lnorm | Classic | 88 (42) | 0.704 [0.591, 0.813] | 117.42 |

| Outcome | Metric | Model | N | R2 | AIC |
| --- | --- | --- | --- | --- | --- |
| T25FW | LN | Whole Brain | 85 | 0.048 | 91.55 |
|  | LV | Whole Brain | 85 | 0.141 | 82.77 |
|  | LN | Tract-based | 85 | 0.141 | 82.78 |
|  | LN | Classic | 85 | 0.134 | 83.50 |
|  | LV | Tract-based | 85 | 0.161 | 80.80 |
|  | LV | Classic | 85 | 0.193 | 77.50 |
|  | Lnorm | Tract-based | 85 | 0.164 | 80.48 |
|  | Lnorm | Classic | 85 | 0.193 | 77.55 |
| 9HPT-D | LN | Whole Brain | 88 | 0.040 | -4.40 |
|  | LV | Whole Brain | 88 | 0.143 | -14.30 |
|  | LN | Tract-based | 88 | 0.229 | -23.64 |
|  | LN | Classic | 88 | 0.183 | -18.53 |
|  | LV | Tract-based | 88 | 0.282 | -29.96 |
|  | LV | Classic | 88 | 0.198 | -20.24 |
|  | Lnorm | Tract-based | 88 | 0.189 | -19.18 |
|  | Lnorm | Classic | 88 | 0.205 | -21.00 |
| 9HPT-ND | LN | Whole Brain | 88 | 0.122 | 23.96 |
|  | LV | Whole Brain | 88 | 0.321 | 1.36 |
|  | LN | Tract-based | 88 | 0.253 | 9.82 |
|  | LN | Classic | 88 | 0.213 | 14.36 |
|  | LV | Tract-based | 88 | 0.379 | -6.51 |
|  | LV | Classic | 88 | 0.359 | -3.73 |
|  | Lnorm | Tract-based | 88 | 0.340 | -1.16 |
|  | Lnorm | Classic | 88 | 0.337 | -0.75 |

